# Functional MRI connectivity accurately distinguishes cases with psychotic disorders from healthy controls, based on cortical features associated with neurodevelopment

**DOI:** 10.1101/19009894

**Authors:** Sarah E Morgan, Jonathan Young, Ameera X Patel, Kirstie J Whitaker, Cristina Scarpazza, Therese van Amelsvoort, Machteld Marcelis, Jim van Os, Gary Donohoe, David Mothersill, Aiden Corvin, Celso Arango, Andrea Mechelli, Martijn van den Heuvel, René S Kahn, Philip McGuire, Michael Brammer, Edward T Bullmore

**Author notes:** SEM and JY contributed equally to the work. MB and ETB contributed equally to the work.

## Abstract

**Background:** Machine learning (ML) can distinguish cases with psychotic disorder from healthy controls based on magnetic resonance imaging (MRI) data, with reported accuracy in the range 60-100%. It is not yet clear which MRI metrics are the most informative for case-control ML.

**Methods:** We analysed multi-modal MRI data from two independent case-control studies of patients with psychotic disorders (cases, N = 65, 28; controls, N = 59, 80) and compared ML accuracy across 5 MRI metrics. Cortical thickness, mean diffusivity and fractional anisotropy were estimated at each of 308 cortical regions, as well as functional and structural connectivity between each pair of regions. Functional connectivity data were also used to classify non-psychotic siblings of cases (N=64) and to distinguish cases from controls in a third independent study (cases, N=67; controls, N = 81).

**Results:** In both principal studies, the most diagnostic metric was fMRI connectivity: the areas under the receiver operating characteristic curve were 92% and 77%, respectively. The cortical map of diagnostic connectivity features was replicable between studies (*r* = 0.31, *P* < 0.001); correlated with replicable case-control differences in fMRI degree centrality, and with prior cortical maps of aerobic glycolysis and adolescent development of functional connectivity; predicted intermediate probabilities of psychosis in siblings; and replicated in the third case-control study.

**Conclusions:** ML most accurately distinguished cases from controls by a replicable pattern of fMRI connectivity features, highlighting abnormal hubness of cortical nodes in an anatomical pattern consistent with the concept of psychosis as a disorder of network development.

## Introduction

Several research studies have used machine learning (ML) algorithms to detect or diagnose psychotic disorders or schizophrenia based on MRI data (1–8). However, in 64 case-control studies of schizophrenia, the ML performance accuracy ranged widely from 59% to 100% (1). Machine learning has been described as a ‘black box’: data go in and a diagnostic decision comes out. It is not always clear which MRI features are most influential diagnostically, and this makes it difficult to validate the machine diagnostic process in terms of what is already known from case-control MRI studies of schizophrenia.

Case-control studies have repeatedly shown that correlations and other symmetric measures of functional connectivity between regional fMRI time series are abnormal in psychotic disorders (9–12). The frontal and temporal cortical hubs of the anatomical connectome have higher probability of grey matter volume deficit in schizophrenia (13), and the association cortical hubs of morphometric similarity networks (from multi-parametric MRI) have abnormal degree, i.e., more or fewer connections to the rest of the brain than in healthy controls, in three independent case-control studies of psychosis (14). The coupling between structural and functional connectivity is reduced in cases of first episode psychosis compared to controls (15). These and other results support the theory that psychosis is a brain network disorder (16–18).

The primary aim of this study was to evaluate the accuracy and replicability of the same machine learning algorithm applied to five different MRI metrics of brain structure and function in case-control studies of psychotic disorders. We considered the functional connectivity matrix between all possible pairs of cortical nodes, estimated from resting state fMRI data; and the structural connectivity matrix, estimated from tractographic analysis of diffusion-weighted imaging (DWI) data. We also considered three local or regional metrics: mean diffusivity (MD) and fractional anisotropy (FA), two DWI markers of cortical microstructure; and cortical thickness (CT), a macrostructural MRI measure. All five metrics were estimated for 308 cortical regions in multimodal MRI data available on participants in two independent case-control studies, collated as part of the PSYSCAN programme (19). Functional MRI connectivity was also evaluated in a third independent, open, case-control study. We predicted that connectivity metrics, compared to local metrics, would support more accurate and replicable ML classification of cases with psychotic disorders.

Our second aim was to ‘open the black box’ of machine learning. Having demonstrated that fMRI connectivity metrics were most informative for ML, we further explored the fMRI connectivity features that were most informative for distinguishing cases from controls based on an individual scan. We compared ML diagnostic feature maps to maps of the group mean differences between cases and controls. In view of evidence that psychotic disorders are related to abnormal brain network development (16–18), we tested the hypothesis that the diagnostic feature map for fMRI connectivity would be co-located with a prior map of aerobic glycolysis, a metabolic pathway that is utilised specifically by neurodevelopmental processes (20), and with a prior map of change in functional connectivity during healthy human adolescence and early adulthood (21). Finally, since psychosis is heritable (22), we tested the hypothesis that fMRI data on non-psychotic siblings of cases would be assigned intermediate probabilities of psychosis by the ML algorithm trained on case and control data.

## Materials and Methods

### Primary studies

We analysed data on three prior case-control studies which had comparable 3T MRI data in one or more MRI modalities for large sample sizes (total N>100 per study). Demographics, questionnaire scores on psychosis symptoms, and measures of MRI quality and head motion, are provided for all three studies in Table 1. All patients satisfied DSM-IV diagnostic criteria for schizophrenia or other non-affective psychotic disorders, and all participants gave informed consent. The studies were ranked Dublin » Cobre » Maastricht in terms of image quality (Table 1 and **SI Table S1**). There were group differences in sex in Maastricht and Dublin (cases were more likely to be male), and a group difference in age in Dublin (cases were older on average). We therefore controlled for age and sex in all subsequent analysis.

**Table 1.** Sample characteristics in three independent case-control studies of psychotic disorder: the Maastricht GROUP dataset (49), the Dublin dataset scanned in the Trinity College Institute of Neuroscience as part of a Science Foundation Ireland-funded neuroimaging genetics study (‘A structural and functional MRI investigation of genetics, cognition and emotion in schizophrenia’), and the Cobre dataset (50). The standing ethics committee of Maastricht University Medical Center approved the Maastricht GROUP study. The St. James Hospital and the Adelaide and Meath Hospital Dublin Incorporating the National Children’s Hospital (AMNCH) joint ethics boards approved the Dublin study. In the Dublin dataset, a different subset of patients had DWI data (82 controls and 33 patients) compared to those with fMRI data (80 controls, 28 patients); see **SI Section S1**.**3**. Medication is given as either AP (on antipsychotic medication), DN (drug naive) or NA (not available). Euler number is a QC metric for MRI data (51): more negative numbers indicate poor image quality. Temporal signal to noise ratio (SNR) is a QC metric for fMRI time series: higher tSNR indicates better quality. Spike percentage and mean framewise displacement (FD) are both measures of head movement during scanning: higher values indicate greater movement effects on fMRI time series (27).

| Dataset | Maastricht |  |  | Dublin |  |  |  | Cobre |  |
| --- | --- | --- | --- | --- | --- | --- | --- | --- | --- |
| Modality | CT/fMRI/MD/FA/DWI |  |  | CT/fMRI |  | MD/FA/DWI |  | fMRI only |  |
| Group | Controls | Cases | Siblings | Controls | Cases | Controls | Cases | Controls | Cases |
| Sample size | 59 | 65 | 64 | 80 | 28 | 82 | 33 | 81 | 67 |
| Age (years) | 29.2 ± 10.3 | 28.1 ± 6.6 | 29.3 ± 8.6 | 28.2 ± 9.0 | 39.9 ± 11.3 | 33.5 ± 12.6 | 42.2 ± 11.7 | 38.9 ± 9.0 | 36.0 ± 12.7 |
| Sex (M) | 23 (39.0%) | 43 (66.2%) | 37 (57.8%) | 38 (47.5%) | 22 (78.6%) | 35 (42.7%) | 24 (72.7%) | 58 (71.6%) | 56 (83.6%) |
| No. of cases with a diagnosis of schizophrenia | N/A | 40 (61.5%) | N/A | N/A | 25 (89.3%) | N/A | 29 (87.9%) | N/A | 58 (86.6%) |
| Medication (AP/DN/NA) | N/A | 63/0/2 | N/A | N/A | 21/0/7 | N/A | 24/1/8 | N/A | 51/2/14 |
| Euler number ± SD | −96 ± 44 | −94 ± 59 | −95 ± 35 | −31 ± 19 | −29 ± 15 | −35 ± 18 | −33 ± 21 | −65 ± 34 | −65 ± 31 |
| Temporal SNR ± SD | 133 ± 23 | 134 ± 22 | 134 ± 19 | 186 ± 27 | 171 ± 24 | N/A | N/A | 164 ± 34 | 165 ± 38 |
| Spike % ± SD | 2.1 ± 1.0 | 2.3 ± 1.1 | 1.8 ± 0.59 | 1.3 ± 0.9 | 2.5 ± 1.5 | N/A | N/A | 2.5 ± 1.6 | 3.1 ± 1.7 |
| Framewise displacement, mean ± SD | 0.19 ± 0.07 | 0.22 ± 0.08 | 0.19 ± 0.06 | 0.22 ± 0.09 | 0.30 ± 0.14 | N/A | N/A | 0.35 ± 0.16 | 0.40 ± 0.17 |
| PANSS positive | N/A | 9.4 ± 3.8 | N/A | N/A | 12.7 ± 2.9 | N/A | 14.5 ± 4.2 | N/A | 15.0 ± 4.7 |
| PANSS negative | N/A | 10.3 ± 5.4 | N/A | N/A | 11.4 ± 5.9 | N/A | 12.5 ± 5.5 | N/A | 15.3 ± 5.5 |

### Macro-structural MRI

T1w MPRAGE images were pre-processed by a prior pipeline (23), using the recon-all (24) command from FreeSurfer (version 6.0). The surfaces were parcellated into 308 cortical regions, using a template derived from the Desikan-Killiany atlas and designed so that regions have similar volumes (23, 25). Cortical thickness (CT) was estimated for each region.

### Micro-structural MRI

Diffusion-weighted images (DWI) were acquired using echo-planar imaging (EPI) sequences with b-value = 1000s*/*mm^2^. Maastricht images had either 76 volumes (43 controls, 44 cases) or 81 volumes (16 controls, 20 cases); Dublin images had 16 volumes. We estimated regional cortical measures of mean diffusivity (MD) and fractional anisotropy (FA) from these images, using FreeSurfer’s trac-all command (26).

### Functional MRI

All fMRI data were acquired using EPI sequences with the following parameters: Maastricht - 200 volumes, acquisition time = 5 mins, 27 slices, TE = 30 ms, TR = 1500 ms, voxel size = 3.5 × 3.5 × 4.0 mm^3^; Dublin – 180 volumes, acquisition time = 6 mins, 35 slices, TE = 30 ms, TR = 2000 ms, voxel size = 3.5 × 3.5 × 3.5 mm^3^; Cobre - 150 volumes, acquisition time = 5 mins, 32 slices, TE = 29 ms, TR = 2000 ms, voxel size = 3 × 3 × 4 mm^3^.

The fMRI images were preprocessed using wavelet despiking (27, 28) as part of a prior pipeline (27), which included slice acquisition correction, rigid-body head motion correction, co-registration to the T1w image, a standard space transform to the MNI152 template in Talairach space, spatial smoothing and intensity normalisation. The time series were band-pass filtered at wavelet scale 2 (29, 30), corresponding to frequency range 0.083-0.17Hz in Maastricht and 0.0625-0.125Hz in Dublin and Cobre. Regions with insufficient signal coverage in one or more participants were excluded from case-control analysis of that study; leaving 248, 290 and 293 cortical regions in the Maastricht, Dublin and Cobre datasets, respectively; see **SI Section S1**.**1**. Group differences in head motion in all three datasets were mitigated by wavelet despiking (27) and regression on 12 motion parameters (translations, rotations and their derivatives); see **SI Section S2**.**3**.

### Functional MRI connectivity and network analysis

Sparse inverse covariance estimation was used to estimate a functional connectivity matrix for each subject, from the regional, normalised (Z-transformed) wavelet coefficients. Sparse inverse covariance is an estimator of partial rather than full correlations (31), meaning it detects only pair-wise specific connectivity between regional nodes, rather than connectivity induced between them by their shared association with a third node (32). The sparse inverse covariance estimator forced the matrices to be symmetric positive definite (SPD), which is advantageous for machine learning.

To calculate degree centrality, we applied a hard threshold to each covariance matrix so that only the top 10% most strongly connected edges were retained in a binary graph. Degree centrality was estimated by the number of edges connecting each node to the rest of the network. For sensitivity analysis, nodal degree was also estimated for graphs thresholded with 5% and 15% connection density; see **SI Section S3**.**7** and **SI Section S7**.

### Structural connectivity and DWI tractography

We corrected DWI images for movement, eddy current distortions and susceptibility distortions. Parcellated T1w MRI images were then registered to DWI images and a diffusion profile was reconstructed for each voxel, using compressed sensing techniques and robust tensor fitting approaches (33), before white matter tracts were identified by deterministic streamline tractography (33). Edges in the DTI networks were weighted by the number of streamlines between each pair of regions and nodal degree was calculated by summing across the rows (or columns) of the DTI connectivity matrix.

### Machine learning

Supervised machine learning algorithms were trained to classify cases and controls, using each of the 5 MRI features in turn. Classification was performed using Gaussian processes (GPs), with a linear covariance function. GPs provide several advantages over more widely used methods such as the support vector machine (SVM) (34) and the linear covariance allows the calculation of feature weights to aid interpretability. For fMRI-based ML, we first took the matrix logarithms of the connectivity matrices, which projects them into Euclidean space (exploiting their SPD property) and has been shown to improve machine classification of fMRI brain networks (35). Age and sex were regressed using a GP based method (36), prior to classification, using the GPML toolbox (37); see **SI Section S3**.**1**.

Consistent with previous studies (38, 39), we performed a 200-fold randomised cross-validation with 10% of subjects allocated to testing and 90% to training, see **SI Section S3**.**4**. We report results in terms of the area under the receiver operating characteristic curve (AUC), leave-one-out (LOO) balanced accuracies, sensitivity, and specificity; see **SI Section S3**.**2**. Per-subject predicted probabilities were calculated as the mean probability of each subject to be classified as a case across all test sets in which that individual was included. The MRI features, e.g. each regional metric or each element of the connectivity matrices, were each associated with a signed ML weight, calculated (34) in each cross validation fold and averaged across folds. For interpretability, fMRI and DTI connectivity matrix feature weights were summed over the edges to obtain regional ML weights.

Finally we performed cross-dataset validation, where we used the Dublin dataset to train the ML classifier for the Maastricht sample, and vice versa, after using the ComBat software (40, 41) to mitigate spurious between-study differences, e.g., arising from different scanning protocols or data quality.

### Case-control analyses

For metrics which followed a Normal distribution (CT, MD, FA), we used a linear regression model, with age, sex and age x sex as covariates, to estimate t-statistics and corresponding P-values for a case-control difference in group means at each region. For functional and anatomical network metrics of nodal degree, which did not follow a Normal distribution, we used a Mann-Whitney U-test to calculate a Z-score and corresponding P-value for case-control difference at each regional node.

We quantified the replicability of the case-control differences in two ways: (i) by correlating the regional case-control t-statistics (or Z-scores) between the Maastricht and Dublin datasets; and (ii) by calculating the number of regions where there was a statistically significant case-control difference after combining the P-values from both datasets using Fisher’s method and applying an FDR correction for multiple comparisons.

### Prior cortical maps of aerobic glycolysis and maturational index

We tested the spatial correlation, or anatomical co-location, of fMRI diagnostic feature maps with two prior cortical maps: a PET-based map of glycolytic index (GI) and an fMRI-based map of adolescent development of functional connectivity (maturational index, −1 < MI < 1) (21). These prior maps were spatially co-registered with the diagnostic feature maps and the correlation between maps was estimated and tested by resampling procedures, including a permutation test controlling for the spatial correlation of metrics in anatomically neighbouring cortical nodes (42).

### Machine detection of MRI data on siblings

To calculate the predicted probabilities of siblings being diagnosed as cases, the GP classifier was trained on the control and patient data and applied to the sibling data. To explore whether siblings also expressed intermediate levels of fMRI connectivity, we used principal component analysis (implemented in MATLAB) to summarise key systems of high (co)variance in the cortical fMRI connectivity profile (248 nodal degrees) for each of the 188 Maastricht participants; then we tested for differences in the first principal component (PC1) between cases, controls and siblings.

### Data and code availability

The code and pre-processed data used in these analyses will be shared via GitHub and FigShare at the time of publication.

## Results

### Diagnostic accuracy of machine learning on five MRI metrics

We trained machine learning classifiers on each of the 5 MRI metrics estimated in each participant in each of the Dublin and Maastricht studies. Receiver operating characteristic curves are plotted for each metric in each study in Figure 1; for more details see **SI Table S5**.

**Fig. 1.**
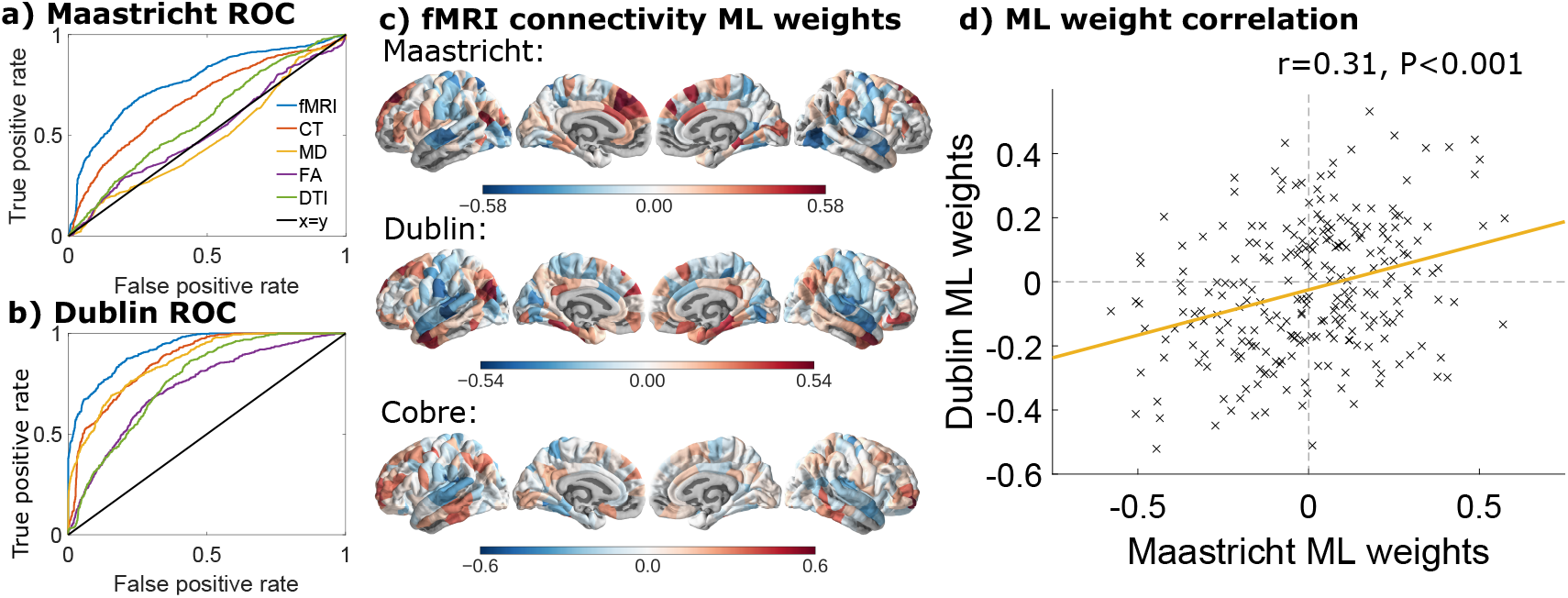
Machine detection of psychotic disorders. Receiver operating characteristic curves (ROCs) for machine learning of diagnosis informed by 5 MRI metrics in (a) Maastricht and (b) Dublin case-control studies of psychotic disorders. An optimal classification will maximise the area under the curve (AUC = 1), so that all patients are correctly diagnosed as cases (true positive rate = 1) when none of the healthy controls are falsely diagnosed as cases (false positive rate = 0). A poor classification will perform at chance, with equal rates of true and false positive diagnosis, and the ROC will follow the (black) line of *y* = *x*. All metrics have closer-to-optimal curves in the Dublin dataset; fMRI connectivity and cortical thickness curves are closer-to-optimal than FA and DWI connectivity curves, in both studies. c) Diagnostic feature maps of the weights assigned to each regional feature for machine diagnosis of psychosis in the Maastricht, Dublin and Cobre datasets. d) Scatterplot of fMRI ML feature weights from Maastricht (*x*-axis) and Dublin (*y*-axis); yellow line is fitted linear regression.

The ML accuracy varied considerably across all 5 metrics and both studies, from ≈ 0.50 to ≈ 0.90. For each metric, ML was more accurate in the Dublin study than in the Maastricht study. The rank order of ML accuracy by different MRI metrics was consistent across the two studies: fMRI connectivity matrices informed the most accurate diagnostic algorithms in both Maastricht and Dublin datasets (AUC= 0.77, LOO= 0.72 in Maastricht; AUC= 0.92, LOO= 0.83 in Dublin), followed by regional cortical thickness (AUC= 0.67, LOO= 0.63 in Maastricht; AUC= 0.87, LOO= 0.76 in Dublin). All other metrics (MD, FA, DWI networks) performed worse in the Maastricht dataset, with accuracies close to chance performance (0.5). In the better quality Dublin dataset (as evidenced by the Euler number and temporal signal to noise ratio (Table 1)), MD informed machine diagnostic accuracy close to the performance informed by CT (AUC ≈0.85). FA and DWI-connectivity performed less well, but still much better than in the Maastricht dataset (AUC ≈ 0.75). Combining the two most predictive MRI metrics (fMRI connectivity matrices and CT) as inputs to the machine classifier did not improve machine diagnostic accuracy compared to using fMRI connectivity matrices alone; see **SI Section S3**.**8**. Predicted probability of caseness was not correlated significantly (*P*_*FDR*_ < 0.05) with psychotic symptom scores, head motion during fMRI, age, or sex; see **SI Section S4**.

### Sensitivity analysis of machine detection based on fMRI connectivity

To further investigate the diagnostic value of fMRI connectivity, we compared ML accuracy based on the full connectivity matrix to ML accuracy based on a 308-length vector of regional degrees. Accuracy remained similar in the Maastricht study (AUC= 0.75; LOO= 0.70); and was somewhat lower in the Dublin study (AUC= 0.85; LOO= 0.76). These results indicate that most of the diagnostic information represented by the full connectivity matrix was captured by the degree centrality or hubness of each node in the fMRI network. For additional independent validation, we also evaluated the ML accuracy in the Cobre dataset based on the fMRI connectivity matrix (AUC= 0.81; LOO= 0.76) or the fMRI network degree vector (AUC= 0.72; LOO= 0.67).

The accuracy of machine detection could be improved for the Maastricht dataset by excluding the patients who did not have a diagnosis of schizophrenia. On this post-hoc subset of 40 patients with schizophrenia, the accuracy increased from AUC= 0.77 (LOO= 0.72) to AUC= 0.81 (LOO= 0.72) for the fMRI connectivity matrix and from AUC= 0.75 (LOO= 0.70) to AUC= 0.77 (LOO= 0.71) for ML based on fMRI degree. Excluding non-symptomatic patients from the Maastricht study also increased the ML accuracy, see **SI Section S3**.**9**.

As a further check for replicability, we performed cross-dataset validation. For the Maastricht sample diagnosed by a Dublin-trained algorithm the AUC was 0.77 (LOO= 0.56); and for the Dublin sample diagnosed by a Maastricht algorithm AUC was 0.76 (LOO = 0.69); see **SI Section S3**.**10**.

### Diagnostic feature maps of machine learning weights

The feature weights from the ML algorithm trained on fMRI data were mapped onto a cortical surface to form a diagnostic feature map for each study; Figure 1. A strong positive ML weight means high fMRI connectivity in that region increased the probability of classification as a case; whereas a strong negative ML weight means low fMRI connectivity in that region increased the probability of psychosis. ML weights were high in association cortex and in the default mode network and low in primary sensory cortex; see **SI Section 8**. The cortical maps of ML weights for fMRI connectivity were significantly correlated between the Dublin and Maastricht studies (*r* = 0.31, *df* = 245, *P* < 0.001). The cortical map of ML weights for the Cobre dataset was also correlated with the corresponding diagnostic feature maps of the Maastricht (*r* = 0.26, *df* = 246, *P* < 0.001) and Dublin (*r* = 0.31, *df* = 280, *P* < 0.001) studies.

Equivalent diagnostic feature maps for CT, MD, FA and DTI connectivity metrics were not significantly correlated between Dublin and Maastricht studies (all *r* < 0.1, *P* > 0.1), except for DWI connectivity which was weakly correlated (*r* = 0.17, *df* = 306, *P* = 0.003); see **SI Figures S4-S7**.

### Replicable case-control differences and machine detection

We estimated case-control group mean differences in each of the 5 metrics for each of the principal studies; see **SI Table S15, SI Table S16** and **SI Section S6**. Normalised difference in fMRI degree (Z-score) was the metric of case-control difference that was most strongly, positively correlated between the Maastricht and Dublin datasets (*r* = 0.62, *df* = 245, *P* < 0.001, Figure 2 a). In both studies, cases had relatively decreased degree in superior temporal and post central cortical areas and increased degree in frontal and inferior parietal cortical areas and the precuneus (Figure 2). Combining P-values over both studies by Fisher’s method, 76 regions showed significant case-control differences in fMRI degree (Figure 2, **SI Table S16**); anatomical details of significantly different regions are provided in **SI Dataset S1**. The case-control fMRI Z-values were also strongly correlated with the diagnostic feature maps for fMRI; for Maastricht *r* = 0.67, *df* = 246, *P* < 0.001, for Dublin *r* = 0.85, *df* = 288, *P* < 0.001.

**Fig. 2.**
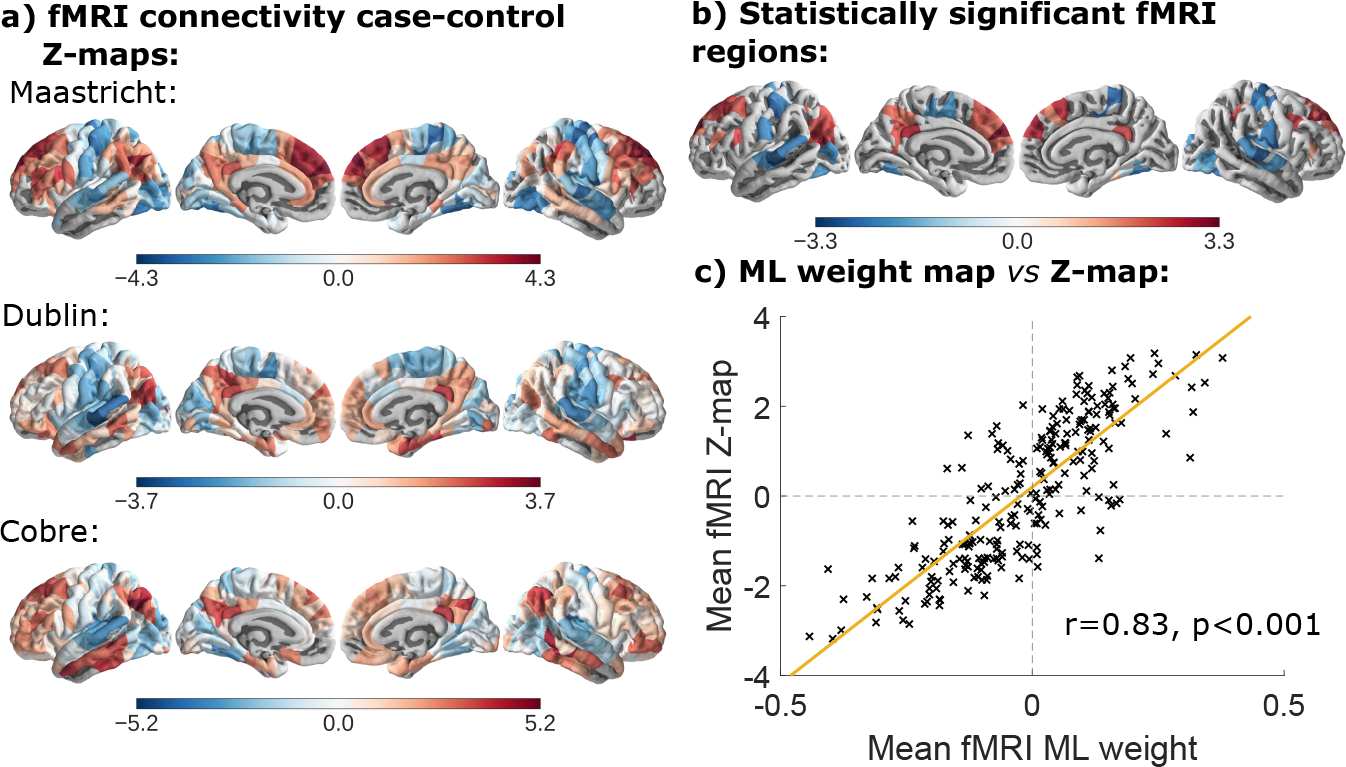
Case-control differences in fMRI degree. a) Z-value maps for the case-control difference in fMRI connectivity strength, or degree, at each cortical node in the Maastricht, Dublin and Cobre datasets. b) The mean Z-value map for nodes with significant case-control difference in fMRI degree defined by combining regional P-values from the Maastricht and Dublin datasets and using the false discovery rate (FDR = 5%) to control for multiple comparisons. c) Scatterplot of Z-values for the mean case-control differences in degree at each regional node, averaged across the Maastricht, Dublin and Cobre studies (y-axis), versus the mean fMRI ML feature weights averaged across the Maastricht, Dublin and Cobre studies (x-axis); yellow line is fitted linear regression.

To confirm the replicability of these results, we estimated cortical case-control Z-values for the Cobre dataset, which were strongly correlated with the corresponding Z-values in the Maastricht and Dublin datasets: *r* = 0.68, *df* = 246, *P* < 0.001 and *r* = 0.60, *df* = 278, *P* < 0.001, respectively. The Cobre case-control Z-values were also strongly correlated with the Cobre diagnostic feature map for fMRI; *r* = 0.74, *df* = 291, *P* < 0.001.

### Neurodevelopmental maps and machine detection

The map of glycolytic index (GI) was negatively correlated with the map of maturational index (MI) (Spearman’s *ρ* = *−*0.54, *P* < 0.001 (21)). The GI map was positively correlated with the fMRI diagnostic feature maps from the Maastricht, Dublin and Cobre datasets, with (*r* = 0.19, *df* = 246, *P* = 0.0025), (*r* = 0.24, *df* = 288, *P* =< 0.001) and (*r* = 0.34, *df* = 291, *P* < 0.001), respectively; see Figure 3. In other words, in regions with high GI, high connectivity increased the probability of classification as a case, whilst in regions with low GI, low connectivity increased the probability of classification as a case. The correlations with the Dublin and Cobre datasets were robust to spatial permutation tests (*P*_*spin*_ < 0.05), although the correlation with the Maastricht dataset was not (*P*_*spin*_ = 0.090).

**Fig. 3.**
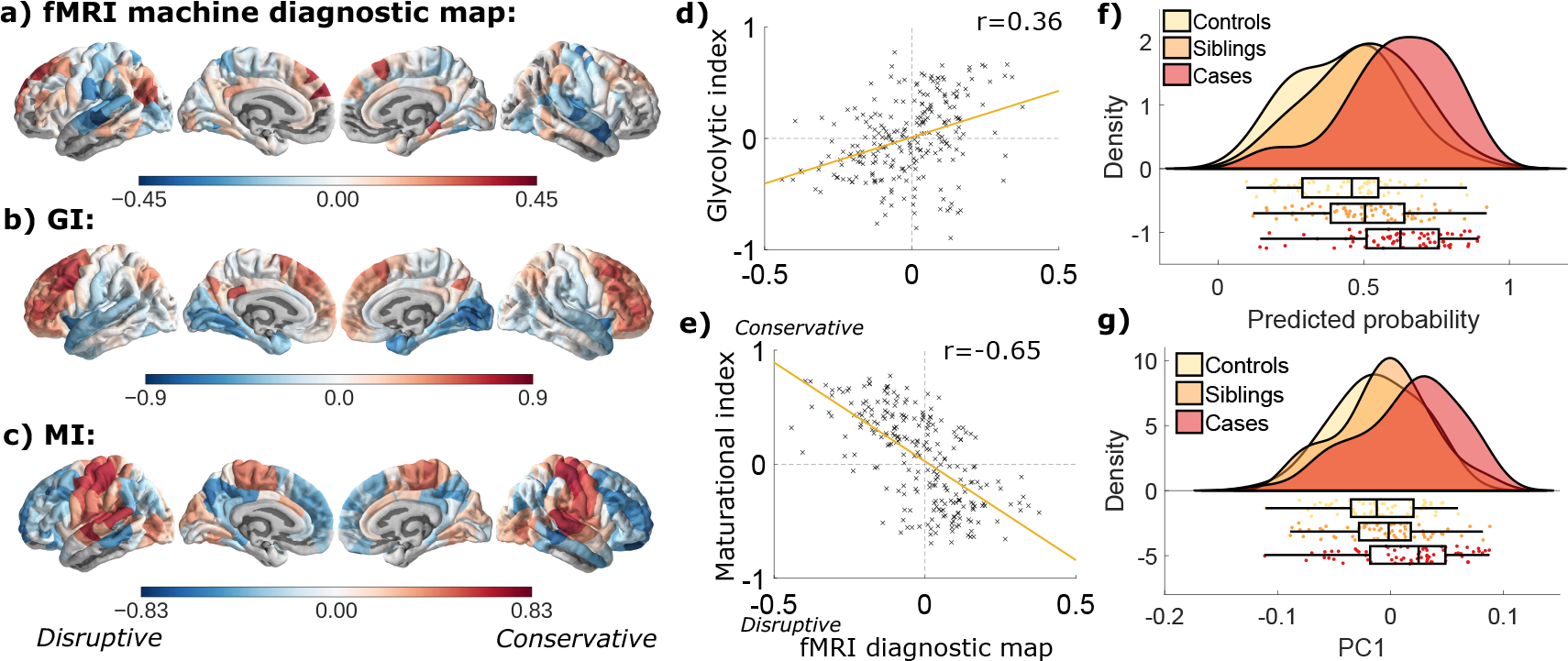
Neurodevelopmental maps, and machine diagnosis of siblings. a) Mean cortical map of psychosis ML weight from fMRI connectivity (averaged across Maastricht, Dublin and Cobre datasets). b) Cortical map of glycolytic index, which measures aerobic glycolysis (20). c) Cortical map of maturational index in healthy adolescence and early adulthood (N=520 fMRI scans, from 298 participants aged 14-26 years) (21). d) Glycolytic index is positively correlated with fMRI diagnostic feature map. e) Maturational index is negatively correlated with the fMRI diagnostic feature map. f) and g): Predicted probabilities of psychosis and PC1 scores (indicative of whole brain functional connectivity) for cases, controls and siblings of cases in the Maastricht dataset. Siblings were assigned intermediate probabilities of psychosis, and had intermediate PC1 scores, compared to cases and controls. Figure created using the RainCloudPlots package (43).

The MI map was negatively correlated with the fMRI diagnostic feature maps from the Maastricht, Dublin and Cobre datasets, with (*r* = −0.42, *df* = 241, *P* < 0.001), (*r* = *−*0.64, *df* = 271, *P* < 0.001) and (*r* = *−*0.38, *df* = 277, *P* < 0.001), respectively; see Figure 3. In other words, in regions with *MI* > 0, located in primary motor and sensory cortex, low fMRI connectivity increased the probability of classification as a case; whereas in regions with *MI* < 0, located in association cortex, high connectivity increased the probability of classification as a case. These results were robust to spatial permutation tests (all *P*_*spin*_ < 0.001).

### Machine detection of siblings

The predicted probability of psychosis for non-psychotic siblings of cases was intermediate between, and significantly different to, the (higher) probability of the cases (two-sided t-test; *P* < 0.001) and the (lower) probability of the controls (two-sided t-test; *P* = 0.04); Figure 3.

Principal component analysis of fMRI connectivity matrices from all participants in the Maastricht study summarised 39% of the total (co)variance in terms of the first principal component (PC1). The cortical map of PC1 loadings is plotted in **SI Figure S12**, and was correlated with the Maastricht fMRI diagnostic feature map (*r* = 0.35, *P* < 0.001). Siblings had PC1 scores that were intermediate between cases and controls, and significantly different to, the (higher) PC1 scores of the cases (two-sided t-test; *P* < 0.001) but not significantly different to the (lower) PC1 scores of the controls; Figure 3.

## Discussion

### Functional MRI informs machine detection of psychosis

Our primary aim was to evaluate how accurately and replicably different MRI metrics could be used to inform machine classification of psychosis. Of the 5 candidate metrics, the most informative was fMRI connectivity, which could be used to classify cases and controls with relatively high accuracy (AUC= 0.92 in Dublin, 0.81 in Cobre, and 0.77 in Maastricht).

This performance is comparable to the highest accuracies previously reported by other large case-control studies (*N* > 100) (1, 5, 44, 45). Importantly, the most diagnostically informative fMRI features, with high positive or negative ML weights, were replicable across three independent studies, with different clinical samples, scanning protocols and geographical locations.

Machine learning accuracies for other MRI metrics were lower, and the diagnostic features were less replicable for structural MRI and DWI metrics. This is a similar pattern to that observed recently (45), which ranked major MRI modalities by classification accuracy for psychosis as fMRI»sMRI»DTI.

### Diagnostic feature maps of psychosis

Our secondary aim was to open the “black box” of machine classification by identifying and contextualising the features that most strongly informed the superior performance of ML based on fMRI connectivity metrics.

First, we found that the machine diagnostic feature maps for fMRI connectivity were strongly correlated with maps of the case-control group mean difference in functional connectivity. This result might seem trivial, however the feature map that decides diagnosis in an individual case is not necessarily co-located with the map of mean differences between cases and controls (1). The high degree of co-location we found suggests that the machine diagnostic process is most likely to classify individuals as cases if their functional connectivity profile is more similar to the abnormal connectivity profile of cases “on average”.

Second, we found that functional connectivity patterns most informative of machine diagnosis of psychosis were co-located with brain systems that are known to be developmentally active during adolescence. The glycolytic index (GI) is a measure of aerobic glycolysis, a metabolic pathway that is utilised specifically by neurodevelopmental processes (20, 46). Maturational index (MI) is a novel measure of adolescent change in functional connectivity measured in a longitudinal fMRI study of healthy young people: *MI* < 0 indicates a region that has “disruptively” increased connectivity during adolescence and early adulthood (14-26 years) from a low baseline level at age 14 (21). Both maps were correlated with the machine diagnostic feature map for psychosis, which is compatible with the theory that psychotic disorders result from aberrant brain network development (16–18).

Third, since psychosis is heritable, and MRI markers have been validated as intermediate phenotypes in studies of non-psychotic siblings (47), we predicted that siblings classified by an ML algorithm trained on case and control fMRI data should be assigned probabilities of psychosis less than the cases but more than the controls. This prediction was supported empirically. Moreover, siblings had intermediate scores on a standard multivariate measure of whole brain functional connectivity (PC1). Taken together, these findings suggest that machine-based detection of psychosis may be sensitive to familial risk of psychosis represented by continuous variation in brain systems of fMRI connectivity.

#### Caveats

The accuracy and between-centre reliability of machine detection of psychotic disorder is not yet good enough, even for the best-performing fMRI connectivity metrics, to be “rolled out” to clinical applications immediately. Improvements in fMRI data acquisition and analysis are expected to improve reliability and accuracy in future. For example, ML performance could be enhanced by a more multivariate and longitudinal approach, combining more, different variables, measured at more than one point in time, and mapping individuals to a more multi-dimensional space than case-control binarization (48). It will also be important to evaluate fMRI-based ML for more clinically critical questions than broad-brush diagnosis of psychotic disorder. For example, how accurately does fMRI connectivity at first episode of psychosis inform ML prediction of clinical outcome?

Case-control designs are vulnerable to the confounding effects of uncontrolled variables and not all the primary studies were well-matched for age and sex. Many other potentially confounding factors, e.g. medication or drug use, were either not measured at all, or not measured consistently across studies. It seems unlikely that the replicable case-control differences reported here are attributable to a replicable but unknown confounding factor that is nothing to do with psychosis. However, more extensive socio-demographic, clinical and cognitive/behavioural data will be important to inform and control fMRI-based ML of psychosis in future.

## Conclusions

Machine learning most accurately and reliably categorised individuals as psychotic disorder cases or healthy controls based on a contextually and theoretically plausible anatomical pattern of dysconnectivity between cortical nodes in resting state fMRI networks. Functional connectomics is a priority candidate for further evaluation as a clinical MRI biomarker of psychosis.

## ACKNOWLEDGEMENTS

This study was supported by grants from the European Commission (PSYSCAN - Translating neuroimaging findings from research into clinical practice; ID: 603196) and the NIHR Cambridge Biomedical Research Centre (Mental Health). The Cobre data was downloaded from the COllaborative Informatics and Neuroimaging Suite Data Exchange tool (COINS; http://coins.mrn.org/dx) and data collection was performed at the Mind Research Network, and funded by a Center of Biomedical Research Excellence (COBRE) grant 5P20RR021938/P20GM103472 from the NIH to Dr. Vince Calhoun. SEM holds a Henslow Fellowship at Lucy Cavendish College, University of Cambridge, funded by the Cambridge Philosophical Society. KJW was funded by an Alan Turing Institute Research Fellowship under EPSRC Research grant TU/A/000017. ETB is supported by a NIHR Senior Investigator Award. The views expressed are those of the author(s) and not necessarily those of the NHS, the NIHR or the Department of Health and Social Care. The LATEXtemplate for this medRxiv submission was created by Ricardo Henriques (and shared under Creative Commons CC BY 4.0).

